# Research on the Demands and Supporting Strategies of Paraeducators for Children with Special Needs in Jiangxi Province from the Perspective of Inclusive Education

**DOI:** 10.64898/2026.08.05.26359305

**Authors:** Zhonglian Fu

## Abstract

Against the backdrop of nationwide inclusive education promotion, children with autism spectrum disorder, intellectual disabilities and other special educational needs (SEN) in Jiangxi Province have raised growing demands for equitable schooling. Paraeducators serve as a critical on-site support mechanism enabling SEN children to access mainstream classrooms; the adequacy of shadow teacher service provision and the maturity of corresponding multi-stakeholder support systems jointly determine the overall quality of local inclusive education. This study adopted mixed quantitative-qualified methods, including questionnaire surveys and semi-structured interviews, to investigate SEN children, paraeducators, general and special education teachers, as well as SEN caregivers across multiple prefecture-level cities in Jiangxi. Grounded in provincial special education policies and local frontline inclusive education practices, we systematically unpacked multidimensional service demands from four core stakeholder groups, diagnosed prominent practical bottlenecks restricting the sustainable operation of shadow teacher services, and constructed regionally tailored multi-layered support strategies aligned with Jiangxi’s educational realities. The findings of this research offer empirical evidence and actionable policy references to advance high-quality inclusive education for SEN children across central China’s Jiangxi Province.

## 1. Introduction

In July 2024, seven central government departments including the China Disabled Persons’ Federation jointly issued the *Implementation Plan for the Autism-Friendly Care Promotion Initiative (2024–2028)*, which explicitly mandates the establishment of shadow teacher systems to facilitate SEN children’s integration into mainstream school settings. The national blueprint *China’s Education Modernization 2035* calls for a sound support system for special education, while the national *14th Five-Year Plan for the Improvement of Special Education Development* proposes phased construction of shadow teacher systems to optimize learning and daily life support for autistic children in mainstream schools. These policy documents establish clear developmental directions for local special education implementation.

As a major educational province in central China, Jiangxi has actively advanced inclusive and equitable special education by incorporating shadow teacher system development into key tasks outlined in its provincial special education upgrading action plans. Designated as a national pilot region for special education reform, Jiangxi has made notable progress in inclusive mainstream placement over recent years yet faces tangible barriers to SEN children’s full integration into general schools. General education teachers typically lack systematic training in special education pedagogy, limiting their capacity to address SEN children’s individualized requirements. For instance, most mainstream instructors cannot fully implement Individualized Education Programs (IEPs) through differentiated homework design, behavioral data collection, or classroom generalization of rehabilitative skills. Due to inherent cognitive, emotional, and behavioral characteristics, SEN children exhibit limited comprehension and compliance with classroom rules, frequently displaying disruptive behaviors such as screaming, physical aggression, and repetitive stereotyped movements among autistic learners. These behaviors disrupt regular teaching routines and hinder independent classroom participation.Paraeducators—professionally trained educational paraprofessionals embedded within mainstream classrooms—effectively mitigate such challenges by providing individualized scaffolding, including verbal prompts and hands-on learning assistance. Their intervention elevates SEN children’s classroom engagement and reduces problematic behaviors, thereby resolving core obstacles to inclusive mainstream placement and safeguarding equal access to school learning and daily activities for SEN children.

Shadow teacher services have already demonstrated practical value in Jiangxi’s inclusive education landscape. Local institutions including Nanchang Xihu Modern Children’s Rehabilitation Center and Nanchang Fuyao Children’s Development Center deploy paraeducators to support autistic children adapting to primary school classrooms. Jiangxi Vocational School of Civil Affairs, in collaboration with relevant administrative departments, has organized public welfare training programs for paraeducators, laying foundational groundwork for the province’s shadow teacher workforce. Nevertheless, field observations reveal pervasive operational challenges within shadow teacher practice. Addressing systemic barriers to shadow teacher support for Jiangxi’s SEN children has emerged as an urgent priority for regional special education development.

## 2. Core Concepts and Theoretical Foundations of Inclusive Education

### 2.1 Definition of Core Concepts

#### 2.1.1 Children with Special Educational Needs

This study defines children with special educational needs as individuals exhibiting physiological, psychological, cognitive, or social-emotional atypicalities requiring targeted special education support, primarily encompassing autistic children, children with developmental delays, students with specific learning difficulties, and other neurodiverse learners.

#### 2.1.2 paraeducators

paraeducators, also referred to as teaching assistants or paraprofessional educators, are specialized practitioners tasked with delivering targeted environmental adaptation support to SEN children within mainstream educational contexts. Their core responsibilities include facilitating classroom engagement, regulating behavioral routines, advancing social communication capacities, and strengthening cognitive and daily living skills for SEN learners.

#### 2.1.3 Inclusive Education

Inclusive education is an educational model that integrates SEN children and neurotypical peers within shared learning environments, with customized supportive services to enable joint learning and holistic development, ultimately advancing educational equity. Rooted in respect for individual differences, inclusive education optimizes educational ecosystems and delivers tailored interventions to meet each SEN child’s developmental demands.

Jiangxi’s inclusive education practice encompasses three primary delivery models: mainstream school inclusive placement, enrollment in specialized special education schools, and home-based outreach instruction. Among these, mainstream inclusive placement constitutes the dominant form, with paraeducators functioning as an indispensable support pillar.

#### 2.1.4 Individualized Supporting Strategies

Individualized supporting strategies denote customized educational intervention frameworks formulated according to each SEN child’s disability classification, developmental level, and personal interests. Adhering to the pedagogy of differentiated instruction, such strategies flexibly adjust intervention modalities and content to match individual developmental gaps, ensuring equitable access to appropriate targeted support for every SEN learner.

### 2.2 Theoretical Foundations

#### 2.2.1 Inclusive Education Theory

Proposed by the United Nations Educational, Scientific and Cultural Organization (UNESCO), inclusive education theory centers on the principle that education must be accessible to all learners regardless of ability level, with every child entitled to equal educational opportunities. The theory rejects segregated special education systems and advocates for co-education alongside neurotypical peers, with schools mandated to provide tailored auxiliary services to accommodate SEN children’s unique needs. This framework delivers the core ideological underpinning for the present study, defining paraeducators’ fundamental mission: facilitating SEN children’s mainstream integration and realizing educational equity, consistent with Jiangxi’s provincial inclusive education developmental orientation.

#### 2.2.2 Applied Behavior Analysis (ABA) Theory

Applied Behavior Analysis constitutes an evidence-based intervention framework grounded in behavioral science. Its core methodology involves systematic functional analysis of SEN children’s behavioral patterns, paired with positive and negative reinforcement techniques to eliminate maladaptive behaviors and cultivate adaptive skills across cognition, social communication, and self-care domains. ABA provides rigorous technical guidance for shadow teacher practice within Jiangxi’s inclusive classrooms, offering standardized interventions for autistic children’s stereotyped movements and aggressive outbursts while building classroom rule awareness and peer interaction competencies—critical to elevating the professional standard of local shadow teacher services.

#### 2.2.3 Situated Learning Theory

Situated learning theory posits that knowledge and skill acquisition occur within authentic contextual environments, through reciprocal interaction between individuals and their surrounding social and physical settings. The theory emphasizes that social communication competence for SEN children must be cultivated in real-world interactive scenarios, with structured simulation of peer exchanges to enable iterative practice of social skills. This framework guides on-site shadow teacher practice across classroom, campus, and household contexts, embedding social skills training within daily routines to strengthen SEN children’s adaptive capacities across diverse social environments.

## 3. Current Demands and Analysis of Shadow Teacher Services for SEN Children in Jiangxi

### 3.1 Research Participants and Research Design

#### 3.1.1 Research Participants

The research subjects of this article include three groups: First, parents of special children. The survey covers a total of 87 children with autism, intellectual disabilities, and speech and language impairments in Nanchang, Jiujiang, Ganzhou, Fuzhou, Yichun, Yingtan, and other cities. Among them, there are 55 children with autism, 16 children with intellectual disabilities, 7 children with learning difficulties, 5 children with speech and language impairments, and 4 children with multiple disabilities. Second, teaching assistants and accompanying readers. A total of 16 teaching assistants and accompanying readers from the aforementioned regions were selected, including full-time teaching assistants, student volunteers, and parent volunteers. Third, teachers. A total of 24 teachers from ordinary schools and special education schools were selected. The research subjects cover different regions and types of groups in Jiangxi Province, ensuring the representativeness and comprehensiveness of the research results.

#### 3.1.2 Research Instruments

This study adopted a mixed-method design combining questionnaires and semi-structured interviews as primary research tools. A self-developed questionnaire titled Questionnaire on the Demand and Supporting Strategies for Paraeducators of Children with Special Needs in Jiangxi Province was used in this research. Initial items were generated via literature review and interviews with frontline practitioners. Content validity was assessed and optimized by five experts in special education, including two associate professors of special education from universities and three principals of special education schools. The average scale-level content validity index (S-CVI/Ave) of the questionnaire reached 0.91.

Preliminary survey data yielded a KMO value of 0.87 with a significant Bartlett’s test of sphericity. Exploratory factor analysis extracted five common factors consistent with the pre-set theoretical dimensions, and all items presented factor loadings above 0.6, indicating satisfactory construct validity. The Cronbach’s α coefficient for the full questionnaire was 0.89, and the α values of each subdimension ranged from 0.76 to 0.85. Thirty participants completed the retest after a two-week interval, with a Pearson correlation coefficient r = 0.84 between the two rounds of scores, demonstrating favorable internal consistency and temporal stability of the instrument.

The questionnaire targeted three groups of respondents: paraeducators, parents of children with special needs, and general education teachers, from whom research data were collected separately. It consists of five sections: (1) basic demographic information; (2) perceptions of demand for paraeducator support; (3) current status and competency requirements of paraeducators; (4) existing paraeducator support systems; and (5) policy and institutional recommendations to improve paraeducator services.

Section 1 collects basic information including respondents’ identities, regional affiliations, and profiles of children with special needs. Section 2 investigates respondents’ understanding of paraeducators’ functions, attitudes toward dedicated paraeducator allocation, core service tasks of paraeducators, and service coverage scope. Section 3 covers the recruitment sources, required professional knowledge, core competencies, and perceived deficiencies of current paraeducators. Section 4 characterizes the present paraeducator support system, including regional policy availability, funding sources for paraeducator services, frequency of professional training, and major operational challenges. Section 5 puts forward targeted suggestions for advancing paraeducator services at governmental and school levels. The complete original questionnaire is provided in Supplementary Material S1.

The semi-structured interview outline was designed for three participant groups: parents of children with special educational needs, school teachers, and paraeducators. The interview guide for parents mainly centered on their demands for paraeducator support, perceived outcomes of paraeducator assistance, as well as school-related requirements and supportive resources. The interview outline for teachers focused on paraeducators’ functions within classroom settings, existing practical challenges, and professional competency standards expected of paraeducators. For paraeducators, the interview questions mainly covered their daily job responsibilities, difficulties encountered during on-site support, intervention effectiveness, and desired institutional and professional support.

A total of nine participants were recruited for the semi-structured interviews, evenly divided into three subgroups with three individuals each: parents of children with special needs, general education inclusive teachers, and paraeducators. All participants were recruited from multiple regions across Jiangxi Province. Among the three parent participants, their children aged 6–10 years, with primary diagnoses of autism spectrum disorder and intellectual disability. The three inclusive teachers provided regular educational support to school-aged children aged 6–12 years, whose primary conditions included autism spectrum disorder, intellectual disability, and specific learning difficulties; all teachers held general normal university degrees without systematic professional training in special education. The three paraeducators also supported children aged 6–12 years, most of whom were diagnosed with autism spectrum disorder. Their educational backgrounds varied: one paraeducator majored in special education, while the other two had non-normal university educational backgrounds. The full original interview outline is provided in Supplementary Material S2.

#### 3.1.3 Research Procedures

##### Field research proceeded in three sequential phases

##### 3.1.3.1. Preparation phase

Relevant academic literature and national/provincial special education policies were systematically reviewed to design standardized questionnaires and semi-structured interview outlines. Research partnerships were established with mainstream inclusive schools, special education schools, and children’s rehabilitation institutions across Jiangxi to confirm participant recruitment pools.

##### 3.1.3.2. Data collection phase

The study was approved by the Ethics Committee of Yuzhang Normal College on October 10, 2025 (approval number: YZNU-EDU-IRB-2025-038). Recruitment of participants began on January 14, 2026, and concluded on April 17, 2026. The survey targets were parents and teachers of children. After obtaining verbal consent, questionnaires and interviews were conducted. Hybrid online-offline surveys and audio-recorded semi-structured interviews were administered, supplemented by on-site observational documentation of shadow teacher classroom practice to collect primary empirical data.

##### 3.1.3.3. Data collation and analytical phase

Questionnaire and interview datasets were coded and thematically analyzed to synthesize core viewpoints, mapping the landscape of unmet service demands and systemic barriers facing shadow teacher provision in Jiangxi.

### 3.2 Core Demands of Children with Special Educational Needs

Jiangxi is currently undergoing a critical developmental phase for inclusive special education. Official 2024 statistics released by the Jiangxi Provincial Department of Education document 37,600 school-aged students with identified special educational needs across the province, generating substantial unmet demand for inclusive auxiliary services to facilitate equal schooling, healthy development, and social integration for neurodiverse learners.

Shadow teacher workforces in Jiangxi comprise special education instructors, university student volunteers, and parent volunteers, with volunteer cohorts constituting the majority and full-time certified practitioners remaining a small minority. Survey data identifies four primary domains of unmet support needs for SEN children accessing mainstream education: individualized academic scaffolding, classroom participation facilitation, emotional and behavioral regulation, and structured social communication training.

**Table 2.** Core Service Demands of Children with Special Educational Needs.

| Service Demand Category | Proportion of Respondents<br>Endorsing Demand |
| --- | --- |
| Classroom academic support (knowledge explanation, differentiated homework guidance) | 60.92% |
| Behavioral intervention and classroom rule-building | 89.66% |
| Social skills training (peer interaction guidance, group activity integration) | 85.06% |
| Daily living skills coaching (dressing, dining, restroom self-care) | 78.16% |
| Home-school communication liaison (daily school progress feedback, relaying parental concerns) | 68.97% |
| Individualized rehabilitative plan implementation (speech therapy, sensory integration training) | 75.86% |

#### 3.2.1 Individualized Tailored Support

Marked heterogeneity in disability type and severity creates divergent individualized support requirements across SEN subpopulations. Autistic children prioritize social communication intervention and emotional behavioral regulation support, requiring paraeducators to mediate peer-teacher communication and mitigate stereotyped or aggressive behaviors to build classroom compliance. Children with intellectual disabilities primarily require structured cognitive and language development interventions, with shadow scaffolding to align learning pace with mainstream curriculum standards. Students with specific learning difficulties demand targeted academic tutoring and metacognitive learning strategy training to enhance knowledge comprehension and homework completion routines. Younger or severely impaired SEN children demonstrate limited comprehension and compliance with classroom and group activity norms, alongside underdeveloped self-care capacities requiring on-site support during mealtimes and restroom routines. Crucially, rehabilitative skills acquired at school (verbal communication, sensory processing) require in-vivo generalization across daily contexts to maximize therapeutic efficacy, a function uniquely delivered by consistent shadow teacher accompaniment.

#### 3.2.2 Classroom Participation Scaffolding

All surveyed SEN children reported consistent need for structured support to enable meaningful classroom participation. Attention deficits, slowed cognitive processing speeds, and executive dysfunction necessitate deconstructed, tiered instructional objectives; for example, segmenting the learning target “word recognition” into three sequential micro-steps: picture-word matching, oral repetition, and independent identification. Impairments to attention, memory, executive function, and emotional regulation frequently prevent SEN children from independently engaging with mainstream instructional activities. paraeducators deliver real-time intervention to redirect inattentive or off-task behaviors (e.g., unscheduled seat departure), sustain task persistence, and support completion of in-class work and take-home assignments.

#### 3.2.3 Emotional and Behavioral Regulation Support

Difficulty regulating emotional arousal and disruptive behavioral outbursts represents a universal challenge for SEN children. Autistic learners commonly exhibit classroom dysregulation including vocal outbursts, desk-tapping, and repetitive stereotyped movements, while children with intellectual disabilities frequently present with tantrums and acute emotional meltdowns. Addressing such incidents requires functional behavioral assessment paired with evidence-based intervention protocols delivered by trained paraeducators. Survey data indicates 91.95% of participating parents reported persistent emotional-behavioral challenges in their children, necessitating dedicated shadow teacher support to cultivate self-regulation and adaptive behavioral repertoires.

#### 3.2.4 Social Communication Support

Social communication impairment constitutes the primary barrier to long-term social integration for SEN children. A combined 85.06% of paraeducators and parent respondents confirmed pervasive social interaction deficits among SEN learners, including limited spontaneous communication with instructors and peers, underdeveloped social etiquette, and chronic social isolation. paraeducators deliver structured social skills coaching to facilitate peer engagement and gradual adaptation to group social environments, addressing a foundational unmet developmental demand for mainstream-integrated SEN children.

### 3.3 Professional and Occupational Demands of paraeducators

#### 3.3.1 Professional Capacity Development Demands

Survey findings document widespread heterogeneity in training and professional backgrounds across Jiangxi’s shadow teacher workforce, dominated by untrained parent-hired paraprofessionals, part-time school instructors, and uncertified volunteers with no formal special education training. A total of 91.95% of survey respondents identified critical gaps in shadow teacher professional competencies, prioritizing training covering special education foundational theory, positive behavioral intervention techniques, individualized education program design, and structured social skills coaching.

#### 3.3.2 Occupational Welfare and Security Demands

The majority of paraeducators work part-time or volunteer without fixed remuneration, while full-time practitioners report low base salaries with minimal social insurance coverage and limited upward career mobility. Virtually all surveyed paraeducators highlighted systemic deficiencies in occupational welfare, citing demand for standardized wage scales, public occupational recognition, comprehensive social security benefits, and formal career advancement pathways, which collectively contribute to low professional self-identity across the workforce.

### 3.4 Institutional and Familial Demands for Shadow Teacher Services

#### 3.4.1 Institutional Demand

Standardized Governance and Resource Allocation Mainstream schools, as primary delivery sites for shadow teacher services, prioritize formalized management frameworks and dedicated inclusive education resource allocation. Survey results reveal most general schools in Jiangxi lack standardized institutional protocols governing shadow teacher practice, with no clear delineation of role responsibilities, operational workflows, or performance evaluation metrics. Critical resource shortages persist across mainstream campuses, including under-resourced special education resource rooms, insufficient rehabilitative equipment, and limited access to full-time certified special education instructors—constraints that undermine shadow teacher service efficacy. Additionally, mainstream general educators lack foundational inclusive education training, creating barriers to coordinated collaborative practice with paraeducators and generating institutional demand for targeted professional development and external supervisory guidance.

#### 3.4.2 Familial Demand: Collaborative Education and Caregiver Training

Parents function as primary long-term caregivers for SEN children, with core unmet needs centering on collaborative home-school intervention and caregiver capacity-building. Survey data indicates 88.51% of participating parents lack formal special education knowledge and evidence-based intervention skills, limiting their capacity to reinforce classroom shadow teacher support within household environments. Caregivers further report barriers to cross-sector resource coordination between schools, rehabilitation clinics, and federated disability services, alongside unmet demand for specialized parenting training and caregiver mental health counseling services.

## 4. Current Landscape and Systemic Dilemmas of Shadow Teacher Support for SEN Children in Jiangxi

### 4.1 Current Status of Shadow Teacher Service Provision in Jiangxi

#### 4.1.1 Dominant Shadow Teacher Service Models

Three primary delivery models of shadow teacher accompaniment operate across Jiangxi’s inclusive education ecosystem:

##### 4.1.1.1. One-to-one individual support

The most widely deployed model, allocated to children with moderate-to-severe disabilities, assigning a dedicated shadow teacher to deliver comprehensive individualized support for a single SEN child; predominantly implemented within mainstream inclusive schools and private rehabilitation clinics.

##### 4.1.1.2. Small-group tiered support

A single shadow teacher supervises cohorts of 3–5 children with mild SEN, delivering targeted group training in cognitive development and social communication; primarily utilized within dedicated special education schools.

##### 4.1.1.3. Whole-class auxiliary support

One shadow teacher provides universal auxiliary classroom support for all SEN children enrolled within a single mainstream classroom; deployed in general schools with low overall SEN enrollment volumes.

Mainstream inclusive placement relies heavily on the rigid one-to-one individual model, lacking differentiated tiered service frameworks tailored to varying disability severity levels, resulting in inflexible, one-size-fits-all service provision.

#### 4.1.2 Workforce Allocation and Professional Competency Status

Two critical workforce constraints persist across Jiangxi’s shadow teacher system: insufficient staffing and widespread under-qualification. In terms of staffing volume, only 18.39% of active paraeducators hold full-time permanent positions, with part-time temporary staff and volunteers comprising the vast majority, generating extreme workforce instability and limited service accessibility for rural and remote SEN families. Regarding professional competency, most shadow practitioners lack systematic pre-service special education training, resulting in inconsistent intervention quality mismatched to heterogeneous SEN children’s individualized needs, frequently escalating home-school conflicts. Two common role misalignment issues emerge in field practice: untrained caregivers adopting a purely custodial “babysitter” role that neglects educational intervention objectives, or overzealous paraprofessionals usurping core instructional decision-making authority from certified classroom teachers. Both misalignments dilute auxiliary service efficacy by misdefining paraeducators’ core supportive function.

#### 4.1.3 Cross-Sector Collaborative Support Status

At the institutional level, most mainstream general schools lack embedded inclusive education frameworks and assign low strategic priority to shadow teacher services, with no formalized cross-stakeholder coordination mechanisms in place. Communication barriers between classroom teachers and paraeducators inhibit integrated educational planning, while a subset of campuses actively resist shadow teacher classroom access, creating operational friction for service delivery.

#### 4.1.4 Provincial Policy and Institutional Support Status

Jiangxi provincial authorities have progressively enacted supportive special education policy frameworks, including the Jiangxi Provincial 14th Five-Year Plan for the Improvement of Special Education Development and the Notice on Strengthening Autistic Children’s Care and Support Services, which explicitly mandate standardized shadow teacher systems and province-wide practitioner training. Despite directional policy guidance, comprehensive regulatory frameworks governing shadow teacher services remain incomplete, characterized by four key gaps:

Absence of standardized entry qualification criteria for shadow teacher practitioners, generating uneven workforce competency;

Fragmented, limited-coverage professional training programs disconnected from on-site practice demands;

Lack of standardized wage compensation and social security protocols for paraeducators;

Missing formalized performance evaluation systems to benchmark and guarantee consistent service quality.

### 4.2 Systemic Dilemmas Facing Shadow Teacher Services in Jiangxi

#### 4.2.1 Occupational Dilemmas Confronting paraeducators

First, ambiguous role delineation represents the primary occupational barrier. Jiangxi has yet to release official regulatory frameworks defining paraeducators’ professional mandate, leading to divergent self-perceptions among practitioners: some frame their role as custodial care providers, while others view themselves as primary classroom instructors, generating inconsistent service focus and diminished intervention outcomes. Second, pervasive professional skill deficits constitute a core operational challenge. As outlined above, most shadow practitioners lack formalized special education training, lacking mastery of functional behavioral assessment, individualized education plan design, and evidence-based intervention methodologies required to deliver differentiated personalized support. Third, chronic low occupational self-efficacy undermines workforce retention. Low societal recognition, inadequate remuneration, and absent career advancement pathways erode professional identity, while persistent misunderstanding from general school teachers and SEN caregivers exacerbates workplace stress and reduces practitioner motivation.

#### 4.2.2 Cross-Sector Support System Dilemmas

Resource scarcity constitutes the primary structural barrier to effective shadow teacher service delivery, compounded by insufficient institutional investment in human, material, and fiscal resources for inclusive education support.

##### 4.2.2.1. Human resource deficits

Understaffed, under-qualified shadow teacher cohorts cannot meet provincial service demand, with severe workforce shortages disproportionately impacting rural remote regions. Mainstream general educators receive minimal formal inclusive education training, while staffing constraints frequently force single paraeducators to simultaneously support multiple SEN children, drastically diluting intervention quality.

##### 4.2.2.2. Material resource deficits

Most mainstream general schools lack fully equipped special education resource rooms and rehabilitative assistive equipment, creating structural barriers to evidence-based shadow teacher practice.

##### 4.2.2.3. Fiscal resource deficits

Dedicated provincial funding streams for shadow teacher remuneration and practitioner training remain underdeveloped and unreliable.

Fragmented cross-stakeholder coordination constitutes a secondary structural barrier. Schools, households, rehabilitation clinics, and disability federations operate in institutional silos with limited formal communication channels, inhibiting cross-system resource sharing and integrated intervention planning.

#### 4.2.3 Policy and Regulatory Dilemmas

Unstandardized workforce entry protocols form the core regulatory gap, with no province-wide minimum competency benchmarks governing shadow teacher recruitment with respect to educational background, clinical intervention skills, and professional ethics. Absent detailed provincial implementation guidelines governing service standards, practitioner certification, and standardized compensation obstruct formalization of a sustainable shadow teacher occupational career track. Low societal awareness and persistent public stigma toward shadow teacher paraprofessional roles further impede recruitment of qualified special education graduates into the workforce. Inadequate occupational welfare frameworks constitute a secondary regulatory barrier: suppressed wage scales and incomplete social insurance coverage generate high workforce turnover, while undefined career ladders eliminate upward professional mobility, severely limiting opportunities to strengthen long-term occupational identity among shadow practitioners. Collectively, these layered systemic barriers create cumulative obstacles for SEN children pursuing inclusive mainstream education, compounded by mainstream teacher capacity constraints, untrained parental intervention practice, and fragmented professional support infrastructure.

**Table 3.**
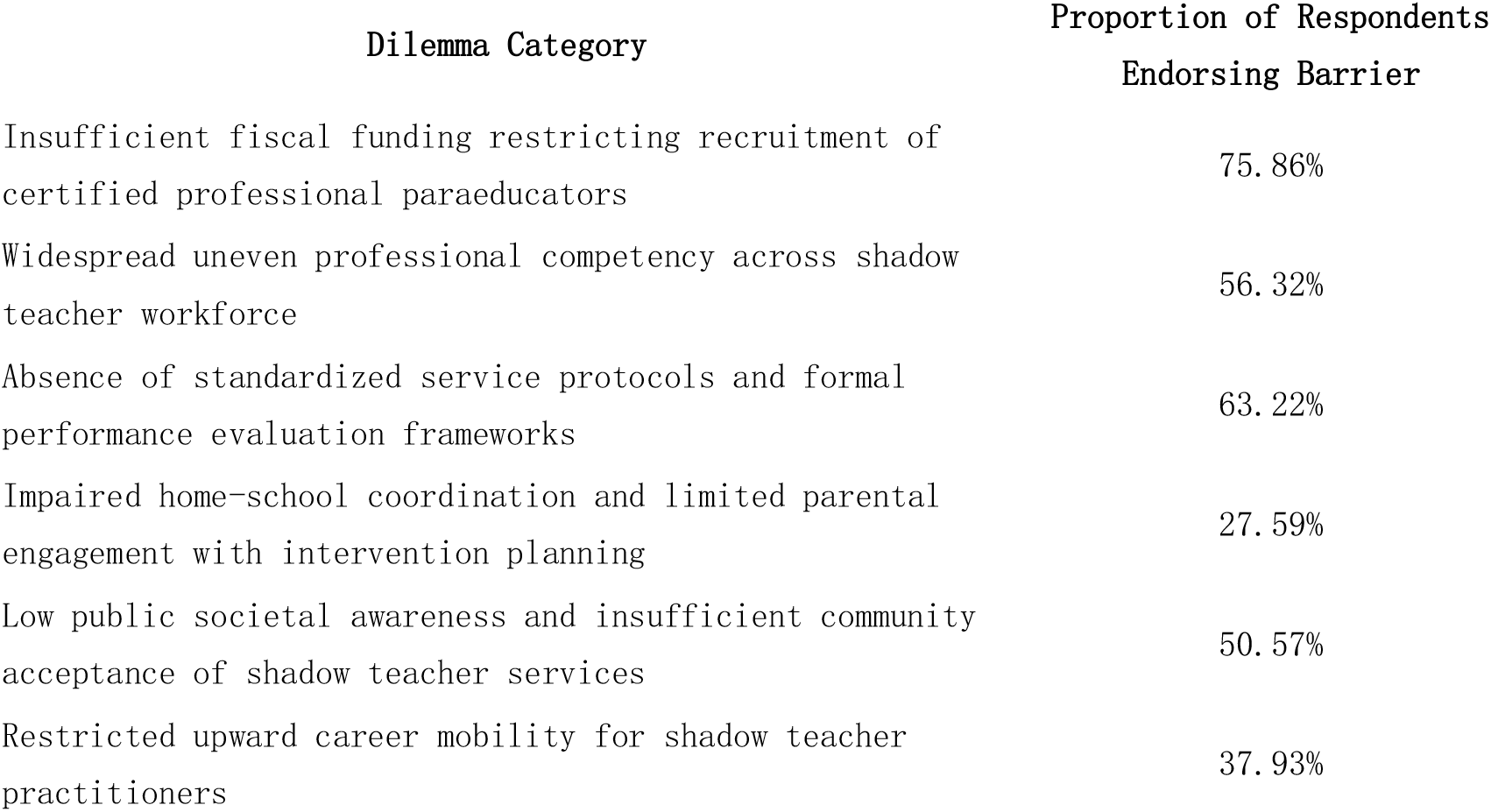
Primary Systemic Dilemmas of Shadow Teacher Services for SEN Children in Jiangxi.

| Dilemma Category | Proportion of Respondents<br>Endorsing Barrier |
| --- | --- |
| Insufficient fiscal funding restricting recruitment of certified professional paraeducators | 75.86% |
| Widespread uneven professional competency across shadow teacher workforce | 56.32% |
| Absence of standardized service protocols and formal performance evaluation frameworks | 63.22% |
| Impaired home-school coordination and limited parental engagement with intervention planning | 27.59% |
| Low public societal awareness and insufficient community acceptance of shadow teacher services | 50.57% |
| Restricted upward career mobility for shadow teacher practitioners | 37.93% |

## 5 Multidimensional Supporting Strategies for Shadow Teacher Services for SEN Children in Jiangxi

Resolving the layered systemic barriers outlined above represents the prerequisite for sustainable shadow teacher service delivery, centered on constructing a four-dimensional integrated support framework coordinated across government, schools, families, and community social sectors. Cross-stakeholder resource integration and targeted actionable interventions form the core roadmap for optimizing Jiangxi’s shadow teacher service infrastructure, systematically strengthening inclusive education provision for provincial SEN children. Survey respondents identified priority government intervention mechanisms as outlined in Table 4, while institutional school-level optimization measures are summarized in Table 5.

**Table 4.**
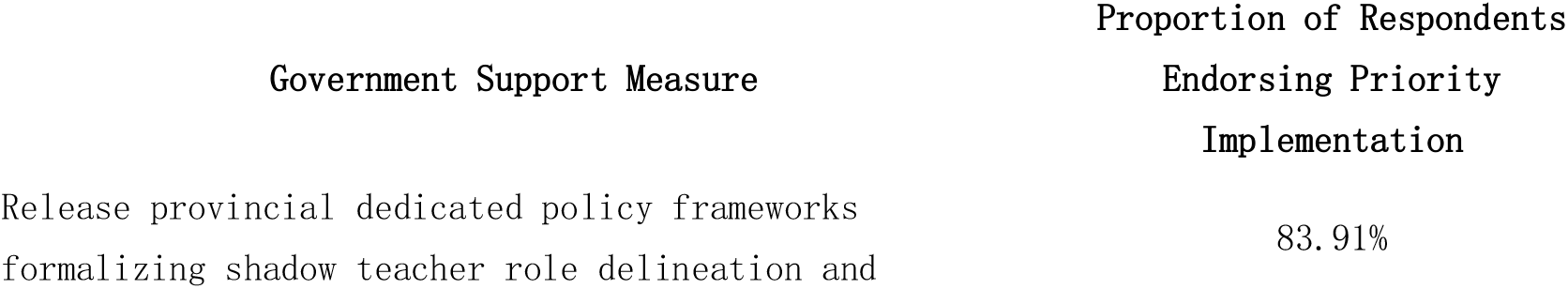
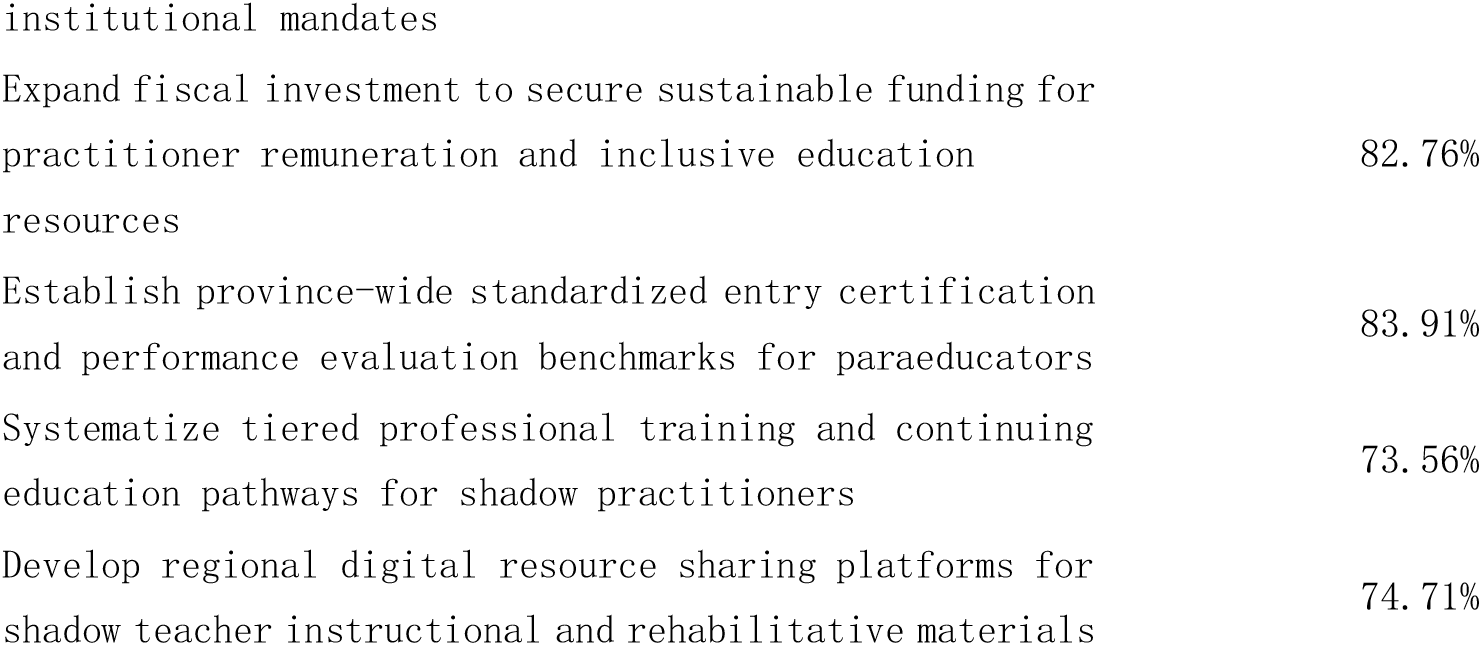
Prioritized Government Interventions to Strengthen Shadow Teacher Service Infrastructure.

**Table 5.** Institutional School-Level Strategies to Optimize Shadow Teacher Service Delivery.

| Institutional Support Measure | Proportion of Respondents<br>Endorsing Priority Implementation |
| --- | --- |
| Formalize clear role delineation between general classroom teachers and embedded paraeducators | 83.91% |
| Establish structured recurring home-school communication protocols for SEN child progress monitoring | 74.71% |
| Deliver on-site instructional supervision and professional mentorship for shadow teacher practitioners | 82.76% |
| Implement standardized periodic outcome evaluations measuring shadow teacher intervention efficacy | 75.86% |
| Cultivate inclusive, accepting campus climates through whole-school awareness-building initiatives | 72.41% |

### 5.1 Government-Level Policy and Regulatory Support

Formalized provincial policy frameworks constitute the foundational structural pillar of sustainable shadow teacher service provision. First, specialized local regulatory instruments must be enacted, including the *Jiangxi Provincial Implementation Measures for Shadow Teacher Services for SEN Children*, which codify standardized role allocation, practitioner qualification thresholds, formal job responsibilities, tiered remuneration schedules, and institutional performance evaluation benchmarks to establish uniform operational guidelines province-wide. Second, sustained provincial fiscal investment mechanisms must be formalized through dedicated annual special education line-item budgets, covering shadow teacher base salaries, system-wide practitioner training programs, and standardized special education resource room construction. Concurrent diversified social financing channels should be cultivated to supplement public fiscal allocations through philanthropic charitable contributions. Third, cross-administrative intersectoral coordination mechanisms must be established to align planning across education, civil affairs, disability federation, health, and human resources authorities, enabling unified provincial strategic planning, cross-agency resource pooling, and standardized service oversight to generate cohesive policy implementation capacity.

Complementary regulatory refinements include drafting the *Jiangxi Provincial Service Guidelines for Special Education Shadow Teacher Support*, establishing formal certification standards, unambiguous role boundaries, and standardized occupational welfare protocols as uniform operational reference documents for all regional service providers. Shadow teacher service infrastructure should be formally integrated into the upcoming *Jiangxi Provincial 15th Five-Year Plan for Special Education Development*, solidifying long-term institutional prioritization. Formal interdepartmental memoranda of understanding will clarify administrative jurisdiction and streamline coordinated policy execution across governing bodies.

### 5.2 Systemic Professional Capacity-Building and Individualized Tiered Service Delivery

Structured professional development constitutes the critical intervention lever to elevate shadow teacher service quality, integrated across government, school, family, and community stakeholder cohorts, with dual priorities of workforce standardization and differentiated individualized support delivery. For shadow teacher workforce development, a three-tiered training certification model spanning provincial standardized training, municipal practitioner credentialing, and school-based day-to-day supervision must be rolled out province-wide. Mandatory certified entry requirements will enforce standardized pre-service training, paired with formalized upward career advancement pathways to strengthen practitioner occupational belonging and professional identity. Concurrently, inclusive education coursework will be incorporated as a mandatory component of all mainstream general educators’ continuing professional development cycles, with recurring specialized training covering special education foundational theory, positive behavioral intervention, and individualized education program co-design. A provincial itinerant special education coaching system will deploy certified special education supervisors to deliver ongoing on-site mentorship to under-resourced mainstream inclusive campuses.

For differentiated individualized service delivery, comprehensive pre-enrollment developmental assessments will be administered for all incoming SEN children to design tailored tiered shadow teacher support frameworks. A graduated fading intervention model will be implemented, transitioning from full one-to-one auxiliary support to partial scaffolded assistance, ultimately progressing toward independent unassisted classroom participation to reduce long-term practitioner reliance. Cross-disciplinary integrated service packages merging academic instruction, clinical rehabilitation, and mental health counseling will be deployed to address the multifaceted developmental demands of SEN children.

Gradient-tiered support frameworks will be institutionalized to match intervention intensity to disability severity: children with severe complex disabilities will receive full-time certified one-to-one shadow teacher support; mild SEN learners will access hybrid collaborative support combining campus special education resource teachers and part-time shadow paraprofessionals; rural and remote SEN children will receive hybrid remote online coaching paired with periodic itinerant on-site shadow teacher services to mitigate regional resource inequities. Leveraging Jiangxi’s provincial university special education departments, designated regional shadow teacher training hubs will deliver standardized core coursework covering neurodiverse child psychology, individualized education planning, and positive behavioral support, formally integrated within the provincial special education resource center’s official teacher training curriculum to systemically elevate workforce clinical competency.

### 5.3 Institutional School-Level Implementation and Campus Inclusive Ecosystem Construction

General schools function as the primary delivery venue for shadow teacher services, requiring targeted institutional intervention across three domains: inclusive physical environment construction, standardized internal governance, and whole-school inclusive cultural cultivation. For inclusive physical infrastructure, campus universal accessibility renovations will be completed province-wide, paired with standardized special education resource rooms equipped with full rehabilitative assistive technology and differentiated instructional materials to support shadow teacher daily intervention practice. For institutional governance, dedicated campus coordinators will be assigned to administer all shadow teacher service operations, formalizing standardized role descriptions and daily operational protocols. Recurring structured communication forums unifying classroom teachers, subject instructors, shadow practitioners, and SEN caregivers will formalize routine home-school progress monitoring. All campuses will implement mandatory collaborative individualized education program (IEP) co-design processes, requiring synchronized intervention planning between general classroom instructors and embedded paraeducators. Shadow teacher performance outcomes will be incorporated into formal school institutional oversight and annual evaluation cycles to guarantee consistent policy implementation.

For inclusive campus cultural cultivation, recurring whole-school inclusive education awareness campaigns will elevate neurotypical students’ and educators’ understanding and acceptance of SEN peers. Structured peer mentorship and cross-group collaborative learning activities will create sustained social integration opportunities for SEN children within a supportive, nondiscriminatory campus environment.

### 5.4 Cross-Sector Community and Family Collaborative Support Network Development

Cross-stakeholder coordination across households and community social sectors constitutes the foundational auxiliary pillar of sustainable shadow teacher service delivery, requiring integrated alignment of familial intervention capacity-building and community-wide support infrastructure. Local universities and specialized children’s rehabilitation institutions will assume primary responsibility for structured shadow teacher vocational training, implementing hybrid theoretical-practical pre-service training pipelines to cultivate a sustained pipeline of qualified professional practitioners. SEN caregivers will access formalized campus-based special education skill workshops and caregiver mental health counseling services to build evidence-based home intervention capacity, fostering balanced collaborative mindsets that avoid over-reliance or outright rejection of shadow teacher auxiliary services and centering family as core complementary intervention partners.

Broader community social support networks will be expanded through formalized volunteer and civil society organization participation in shadow teacher service provision to expand regional workforce capacity. Mainstream media outreach will disseminate inclusive education messaging to elevate public societal awareness and reduce occupational stigma surrounding shadow paraprofessional roles. Community-based family support hubs will facilitate cross-household resource linkage, peer caregiver mutual aid, and cross-sector service coordination to build a cohesive integrated regional support ecosystem.

Jiangxi’s provincial pilot implementation of standardized shadow teacher services for SEN children demonstrates the province’s institutional commitment to advancing educational equity and safeguarding the developmental rights of neurodiverse vulnerable youth.

Systematic dissemination of the empirical findings and policy recommendations outlined within this study will support the incremental maturation of a comprehensive,multi-stakeholder shadow teacher support framework for inclusive education across Jiangxi Province.

## Supporting information

Supplemental material 1 will be used for the link to the file on the preprint site.

Supplement material 2 will be used for the link to the file on the preprint site.

## Data Availability

All data produced in the present study are available upon reasonable request to the corresponding author.

https://figshare.com

