## Supplemental material 1 will be used for the link to the file on the preprint site. for "Research on the Demands and Supporting Strategies of Paraeducators for Children with Special Needs in Jiangxi Province from the Perspective of Inclusive Education"

### Introduction

Dear respondents,

This questionnaire aims to investigate the practical demands and existing support status of paraeducator services for children with special needs in Jiangxi Province. The findings will provide empirical evidence to optimize educational and rehabilitation services and improve the support system for paraeducators. All responses are anonymous, and the collected data will only be used for academic research. Please answer all items truthfully based on your real experience. Thank you sincerely for your participation and support!

### Part 1 Basic Information

1. Your identity:

☐ Parent/guardian of a child with special needs
☐ Special education school teacher
☐ General inclusive classroom teacher
☐ Paraeducator
☐ Administrator of education authority
☐ Others (please specify: ________)

2. Your region within Jiangxi Province:

☐ Nanchang ☐ Jiujiang ☐ Ganzhou ☐ Shangrao ☐ Yichun ☐ Ji’an ☐ Fuzhou ☐ Xinyu ☐ Pingxiang ☐ Yingtan ☐ Jingdezhen

3. If you are a parent/guardian of a child with special needs, please fill in your child’s information: (1) Child’s age: ______ years old
(2) Child’s disability type: ☐ Autism spectrum disorder ☐ Intellectual disability ☐ Physical impairment ☐ Visual impairment ☐ Hearing impairment ☐ Speech and language disorder ☐ Emotional and behavioral disorder ☐ Multiple disabilities ☐ Others (please specify: ________)
(3) Child’s education placement: ☐ Full-time special education school ☐ Inclusive general classroom ☐ Home-based outreach teaching ☐ Homeschooling

4. If you are a teacher or paraeducator, your years of working experience:

☐ 1 year or less ☐ 2 - 5 years ☐ 6 - 10 years ☐ Over 10 years

### Part 2 Perceptions of Demands for Paraeducator Support

1. How well do you understand the definition and functions of paraeducator services? ☐ Not at all ☐ Slightly ☐ Moderately ☐ Quite well ☐ Extremely well

2. Do you think children with special needs should be assigned dedicated paraeducators? ☐ Strongly disagree ☐ Disagree ☐ Neutral ☐ Agree ☐ Strongly agree

3. If you agree that paraeducators are necessary, which core services do you expect them to provide? (Multiple choices allowed)

☐ In-class academic support (knowledge explanation, homework guidance)
☐ Behavioral intervention and management (correct challenging behaviors, build routine awareness)
☐ Social skill development (facilitate peer interaction and group integration)
☐ Daily living skills training (dressing, dining, toilet training, etc.)
☐ Home-school communication bridge (report school performance, transmit parental requests)
☐ Implementation of individualized rehabilitation plans (speech therapy, sensory integration training)
☐ Others (please specify: ________)

4. Which time periods should paraeducator services cover? (Multiple choices allowed)

☐ Formal class hours
☐ Recess breaks
☐ Lunch and noon rest time
☐ Extracurricular/physical education sessions
☐ Before/after-school drop-off and pick-up
☐ Others (please specify: ________)

5. What student-to-paraeducator ratio do you consider appropriate?

☐ 1:1 (one paraeducator for one child)
☐ 1:2 (one paraeducator for two children)
☐ 1:3 (one paraeducator for three children)
☐ Flexible allocation (adjusted according to children’s disability severity)

### Part 3 Current Status and Competency Requirements of Paraeducators

1. What are the main sources of paraeducators you have encountered? (Multiple choices allowed)

☐ Full-time school teachers with additional paraeducator duties

☐ Professionals recruited via government service procurement

☐ Volunteers from non-profit organizations

☐ Paraeducators privately hired by parents

☐ Student interns majoring in special education from universities

☐ No paraeducator available at present

☐ Others (please specify: ________)

2.What professional knowledge should a qualified paraeducator master? (Multiple choices allowed)

☐ Theories of special education

☐ Child developmental psychology

☐ Rehabilitation training methods for different disabilities

☐ Behavioral intervention techniques (e.g., ABA, positive behavioral support)

☐ Classroom organization skills

☐ First aid and emergency response capabilities

☐ Home-school communication skills

☐ Others (please specify: ________)

3.What core competencies should paraeducators possess? (Multiple choices allowed)

☐ Observational and assessment skills (identify children's needs and behavioral challenges)

☐ Ability to design and implement individualized support plans

☐ Emotional regulation skills (manage children's emotional outbursts)

☐ Teamwork skills (collaborate with teachers and families)

☐ Stress tolerance

☐ Others (please specify: ________)

4.What is the most prominent deficiency among current paraeducators? (Multiple choices allowed)

☐ Theoretical professional knowledge

☐ Practical intervention skills

☐ Communication and collaboration abilities

☐ Professional identity and sense of belonging

☐ Emergency response capacity

☐ Others (please specify: ________)

### Part 4 Current Status of the Paraeducator Support System

1.Does your local region have clear policy documents supporting paraeducator services?

☐ Complete formal policies

☐ Preliminary policy guidelines

☐ No relevant policies

☐ Unclear

2.What are the primary funding sources for paraeducator services? (Multiple choices allowed)

☐ Government fiscal appropriation

☐ School operating funds

☐ Full payment by children’ s parents

☐ Donations from social welfare organizations

☐ No financial support available

☐ Unclear

3.Do local paraeducators receive regular professional training?

☐ Monthly or more frequent

☐ Quarterly

☐ Annually

☐ No training at all

☐ Unclear

4.What major challenges exist for paraeducator services at present? (Multiple choices allowed)

☐ Insufficient funding to hire qualified professionals

☐ Uneven professional competency among practitioners

☐ Lack of unified service standards and evaluation mechanisms

☐ Poor home-school cooperation and low parental engagement

☐ Low public awareness and insufficient social recognition

☐ Limited career development pathways for paraeducators

☐ Others (please specify: ________)

### Part 5 Suggestions for Optimizing Paraeducator Support Systems

1.What measures should governments take to advance paraeducator services? (Multiple choices allowed)

☐ Issue special policies to clarify paraeducators’ roles and responsibilities

☐ Increase fiscal investment to secure service funds and staff salaries

☐ Establish unified entry criteria and assessment standards for paraeducators

☐ Organize systematic professional training and continuing education ☐ Build regional resource-sharing platforms for paraeducators ☐ Others (please specify: ________)

2.How can schools optimize paraeducator work arrangements? (Multiple choices allowed)

☐ Clarify the division of responsibilities between teachers and paraeducators

☐ Set up regular home-school communication routines

☐ Provide on-site guidance and support for paraeducators

☐ Conduct regular evaluations of paraeducator service outcomes

☐ Foster an inclusive and accepting campus culture

☐ Others (please specify: ________)

3.How should parents cooperate with paraeducators? (Multiple choices allowed)

☐ Proactively share children' s family situations and personalized needs

☐ Participate actively in home-school collaborative activities

☐ Cooperate with paraeducators to implement home rehabilitation tasks ☐ Respect paraeducators' work and build mutual trust

☐ Others (please specify: ________)

4.Do you have any additional recommendations to improve the paraeducator support system for children with special needs in Jiangxi Province?
