## Supplement material 2 will be used for the link to the file on the preprint site. for "Research on the Demands and Supporting Strategies of Paraeducators for Children with Special Needs in Jiangxi Province from the Perspective of Inclusive Education"

Supplementary Material S2:Semi-structured Interview Guides

#### Interview Guide for Parents of Children with Special Needs

Hello. I am a university researcher focusing on the education and rehabilitation of children with special needs. This interview aims to investigate the current situation of paraeducator services for children with special needs in Jiangxi Province, and we sincerely appreciate your participation. The whole conversation will be audio-recorded solely for research data collection. All interview content will be anonymized in subsequent analysis and manuscripts, and your personal information will not be disclosed. Thank you for your understanding and support.

##### Interview Questions

1. What type of disability does your child have, and how old is your child?
2. Do you think your child needs a dedicated paraeducator? Please explain your reasons for or against this need.
3. In your opinion, what is the core purpose of paraeducator support?
4. What daily tasks do you believe paraeducators should undertake?
5. What positive impacts do you think paraeducators bring to children’s development?
6. Does paraeducator support exert any influence on you and your family? If yes, please elaborate.
7. Who do you believe should take primary responsibility for children’s academic learning and daily life at school?
8. Have you ever accompanied your child at school as a caregiver? If so, what specific work did you complete, and what difficulties did you encounter?
9. Does your child have a dedicated paraeducator? Are you satisfied with the outcomes of their support?
10. Does the school set formal requirements for paraeducators? If yes, what specific regulations are in place?
11. Does the school provide professional training opportunities for paraeducators?
12. What kinds of support and assistance does the school offer to paraeducators?

#### Interview Guide for General Inclusive Teachers

Hello. I am a university researcher focusing on the education and rehabilitation of children with special needs. This interview aims to investigate the current situation of paraeducator services for children with special needs in Jiangxi Province, and we sincerely appreciate your participation. The whole conversation will be audio-recorded solely for research data collection. All interview content will be anonymized in subsequent analysis and manuscripts, and your personal information will not be disclosed. Thank you for your understanding and support.

##### Interview Questions

1. Do you think paraeducator support is necessary within school settings? Please explain your reasons for or against this arrangement.
2. In your opinion, what is the core purpose of paraeducator support?
3. What daily tasks do you believe paraeducators should undertake?
4. What positive impacts do you think paraeducators bring to children’s development?
5. Does the presence of paraeducators affect your daily teaching work? If yes, please elaborate.
6. Who do you believe should take primary responsibility for students’ academic learning and daily life at school?
7. What specific work do paraeducators actually perform during school hours?
8. What strengths do paraeducators demonstrate in their work? What challenges do they commonly face?
9. Will you take the initiative to communicate your views on paraeducator support with paraeducators?
10. Will you accept and consider suggestions about students provided by paraeducators?
11. Are you satisfied with the overall effectiveness of current paraeducator services?
12. Does the school set formal requirements for paraeducators? If yes, what specific regulations are in place?
13. What kinds of support and assistance does the school offer to paraeducators?
14. Are you satisfied with the school’s current support for paraeducators?
15. What additional institutional support and resources do you believe paraeducators should receive from schools?

### Interview Guide for Paraeducators

Hello. I am a university researcher focusing on the education and rehabilitation of children with special needs. This interview aims to investigate the current situation of paraeducator services for children with special needs in Jiangxi Province, and we sincerely appreciate your participation. The whole conversation will be audio-recorded solely for research data collection. All interview content will be anonymized in subsequent analysis and manuscripts, and your personal information will not be disclosed. Thank you for your understanding and support.

#### Interview Questions

1. How long have you worked as a paraeducator?
2. What types of children with special needs have you supported, and what is their age range?
3. What is your highest educational attainment and major? Do you have any academic background related to special education?
4. What is your monthly income as a paraeducator?
5. What are your general perceptions of children with special needs?
6. What core tasks do you complete during daily paraeducator work, and what major difficulties have you encountered?
7. What influences does your support bring to the children you assist?
8. Do you receive adequate support and assistance from the school during your work?
9. Are you satisfied with the support provided by the school?
10. Are you satisfied with the cooperation and support you receive from children's parents?
11. What additional types of support and resources do you hope to obtain in your work?
